# Cortical motor network flexibility during lower limb motor activity and deficiencies after stroke

**DOI:** 10.1101/2020.06.15.20130773

**Authors:** Jacqueline A. Palmer, Trisha M. Kesar, Steven L. Wolf, Michael R. Borich

## Abstract

**Background and Purpose:** Motor network flexibility, the ability to switch between different neural network connectivity configurations, is a key feature of normal motor behavior and motor learning. Limited flexibility of the motor system to adapt to environmental stimuli is a pervasive feature of mobility impairment in chronic stroke survivors. The reduced capacity to modulate cortical motor network connectivity during motor behavior has been implicated as a key neural mechanism for constrained flexibility of the motor system. However, whether the theory of reduced cortical motor network flexibility prevails in the lower limb corticomotor system and in the context of functional lower limb motor behaviors remains unknown. In this study, we tested the capacity of the lower limb motor cortex to react to external stimuli using transcranial magnetic stimulation (TMS) and the ability of the cortex to modulate motor network connectivity during lower limb motor activity in a group of chronic stroke survivors and age-matched older adult controls. We evaluated the relationship between activity-dependent modulation of TMS-evoked motor cortical connectivity and post-stroke clinical and biomechanical walking function as well as corticospinal excitability of the paretic leg.

**Methods:** Chronic stroke survivors (n=14) and older adults (n=9) completed concurrent TMS-electroencephalography (EEG) testing, clinical evaluation, and biomechanical walking assessment. EEG was recorded during TMS delivered over the contralesional (c)M1 and ipsilesional (i)M1 during rest and active ipsilateral plantarflexion. Interhemispheric connectivity was calculated as the post-TMS (0-300ms) imaginary part of coherence value between electrodes overlying cM1 and iM1 within the beta frequency range (15-30Hz). We compared cM1 and iM1 TMS-evoked beta coherence between groups during rest and active conditions and tested associations with walking impairment and paretic leg corticospinal excitability in stroke survivors.

**Results:** In older adult controls, TMS-evoked beta coherence was greater compared to stroke survivors and showed a reduction from rest during active motor conditions regardless of hemisphere. Stroke survivors showed lower TMS-evoked beta coherence during cM1 TMS and a lack of modulation between rest and active conditions. Higher cM1 TMS-evoked beta coherence at rest and reduction of beta coherence during paretic plantarflexion was associated with greater paretic ankle moment during walking and lower levels of clinical lower limb motor impairment. Increased beta coherence during iM1 TMS at rest was associated with the presence of a MEP in the paretic lower limb.

**Conclusions:** We found that, like findings in animal models and the upper limb, the lower limb corticomotor system showed reduced cortical network reactivity to external stimuli (TMS), and impaired modulation of cortical network connectivity during lower limb motor activity after stroke. Impaired cortical reactivity and modulation were associated with post-stroke clinical and biomechanical walking function and corticomotor excitability of the paretic leg, supporting the link between cortical network connectivity, motor network flexibility and lower limb motor behavior. These findings have important implications for the development of targeted and individualized treatments to improve lower limb disability in stroke survivors.

## Introduction

The ability to switch between different neural network connectivity configurations is a key feature of normal motor behavior and motor learning.^1,2^ This motor network flexibility can be indexed by functional cortical connectivity, which is characterized by dynamic modulation between rest and motor activity^1^ and reactiveness to external stimuli (**Figure 1**).^3^ After stroke, impaired modulation of motor cortical network connectivity and reduced cortical reactivity to transcranial magnetic stimulation (TMS) have been observed during paretic upper limb motor activity.^1,4–6^ Impaired cortical connectivity modulation and reactivity were also associated with greater paretic hand impairment, suggesting a supportive role of motor network flexibility in post-stroke upper limb motor function.^1,4^ In the context of lower limb motor behaviors, motor network flexibility may be important to dynamically modify movement patterns during constantly changing environmental demands.^7^ Indeed, stroke survivors show not only impaired lower limb motor control but also reduced variability of movement patterns during walking and standing balance.^7,8^ These observations suggest motor network flexibility may support functional mobility and lower limb motor recovery after stroke. If so, understanding motor network flexibility may have important implications for post-stroke rehabilitation because higher levels of motor pattern variability have been linked to faster rates of motor learning and can be modulated with training.^9^ Despite considerable interest in motor network flexibility as it relates to motor learning and upper limb recovery after stroke, little is known about the neural origins of motor network flexibility in the lower limb motor system or its potential role in post-stroke functional lower limb motor behaviors such as walking.

**Figure 1.**
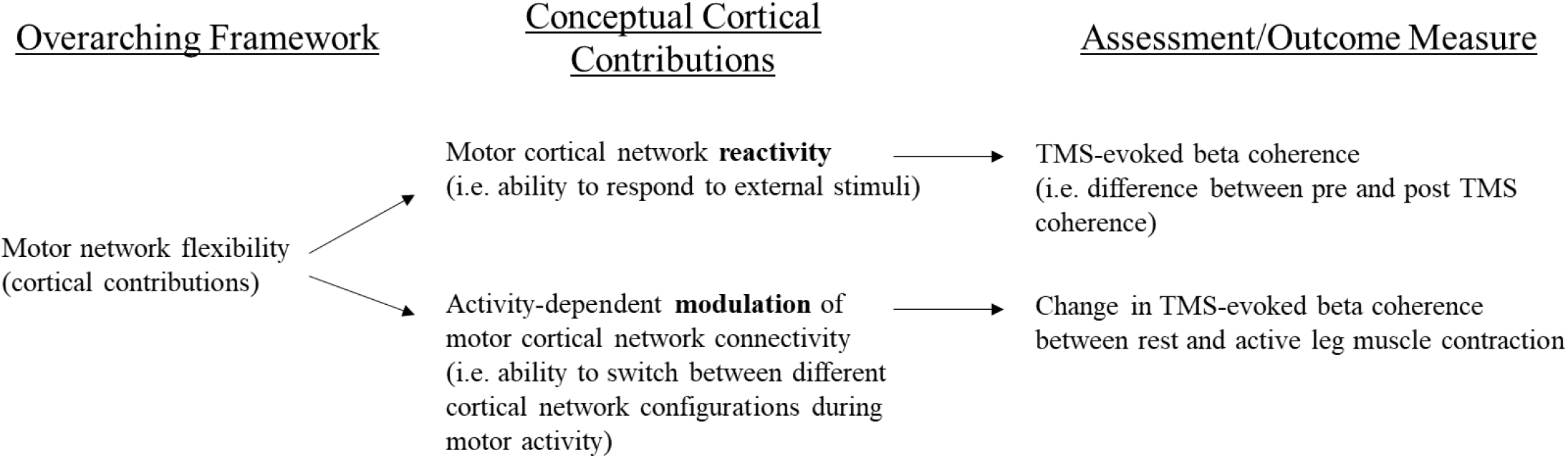
Conceptual framework and assessment of cortical contributions to motor network flexibility of the lower limb corticomotor system.

Modulation of cortical network connectivity in the beta frequency band (15-30Hz) is thought to reflect γ-aminobutyric acid (GABA) neural network interactions linked to sensorimotor activity.^10–13^ Typically, cortical beta oscillations are attenuated during voluntary movements.^14,15^ A lack of normal modulation of beta activity during movement has been observed after stroke,^16^ and may reflect deficient motor network flexibility.^17^ In this theory, the clinical manifestations of the inability to “switch” from one neural network configuration to another are observed in the impaired generation of new movement patterns and the inability to effectively respond to environmental stimuli.^18,19^ This evidence raises the possibility that impaired modulation of cortical beta network connectivity may contribute to pervasive features of post-stroke lower limb hemiparesis, particularly the inability of the paretic leg to generate propulsive forces.^20,21^

The reactive dynamics of functional cortical connectivity to external stimuli can be probed directly using a concurrent TMS-electroencephalography (EEG) approach.^22,23^ The utility of TMS-evoked cortical responses may circumvent some previous technical challenges of assessing lower limb motor cortical connectivity,^24,25^ and can be particularly useful in clinical populations with reduced corticospinal tract integrity.^3^ Additionally, TMS-evoked cortical responses can be measured in task-based paradigms on millisecond timescales^3^ to study task-related information processing during motor activity.^4^ Variability of TMS motor evoked potentials (MEPs), an inherent feature of TMS-electromyography (EMG) measures, appear to reflect motor network flexibility that has been positively linked to motor learning.^26^ Recent studies demonstrated that dynamic cortical oscillatory activity could explain MEP variability,^27–29^ providing insight into the dynamic cortical mechanisms underpinning motor network flexibility. Still, lower limb motor network flexibility may not have a cortical origin, as subcortical networks substantially contribute to mobility function and, in some cases, recovery of mobility after stroke.^30^ Direct measures of TMS-evoked cortical connectivity in lower limb motor regions could help elucidate the neural origins and mechanisms underpinning lower limb motor network flexibility.

Impaired interhemispheric motor cortical network connectivity may be a specific attribute of attenuated motor network flexibility in chronic stroke survivors.^1,6,31^ Recently, we observed that in response to ipsilesional primary motor cortical (iM1) TMS, stroke survivors showed reduced cortical reactivity and modulation of interhemispheric beta connectivity between upper limb motor regions.^4^ Individuals showing greater TMS-evoked interhemispheric connectivity during motor activity demonstrated better paretic hand function, supporting the positive role of interhemispheric cortical motor network flexibility to post-stroke upper limb function.^4^ Interhemispheric connectivity may be particularly salient to functional lower limb behaviors, such as walking and standing balance, where the behavior of one limb directly affects that of the contralateral limb.^32^ However the neural underpinnings of flexible interlimb coordination necessary for functional lower limb motor behaviors remain unclear.^25^ Previous studies have observed that nonparetic leg motor activity influences TMS-evoked MEPs and motor performance in the paretic leg.^33–36^ A possible explanation for neuromotor interference between limbs is that impaired activity-dependent interhemispheric connectivity modulation between motor cortices may lead to impaired lower limb motor control after stroke.

In this study, we investigated motor network flexibility of the lower limb corticomotor system by assessing interhemispheric cortical connectivity between lower limb motor regions. We evaluated TMS-evoked cortical reactivity and modulation of TMS-evoked cortical connectivity during leg motor activity in a group neurologically intact older adults and stroke survivors (**Figure 1**). We tested the behavioral significance of motor network flexibility to paretic leg motor impairment and post-stroke walking dysfunction. We also tested the association between TMS-evoked cortical reactivity and the ability to elicit MEPs in the paretic lower limb.

## Methods

Fourteen individuals (age: 66 ± 11years, 4 females) with chronic stroke (>6 mo.) and nine neurologically intact older adults (age: 68 ± 6 years, 3 females) participated in this study (**Table 1**). All participants completed a single neurophysiologic testing session. On a separate day, a subset of 11 stroke participants completed an additional testing session for clinical and biomechanical walking assessment. Inclusion criteria for participants in the stroke group included a single ischemic cortical or subcortical stroke confirmed by magnetic resonance imaging (MRI) and the ability to walk at least 10 meters without the assistance of another person. Participants were excluded if they had hemorrhagic stroke, a stroke directly affecting the lower extremity region of M1 or the corpus callosum, history of multiple strokes, joint contractures affecting the lower extremity joints, neurodegenerative or orthopedic disorder or psychiatric diagnosis, or contraindications to TMS.^37^ The experimental protocol was approved by the Emory University Institutional Review Board and all participants provided written informed consent.

**Table 1.**
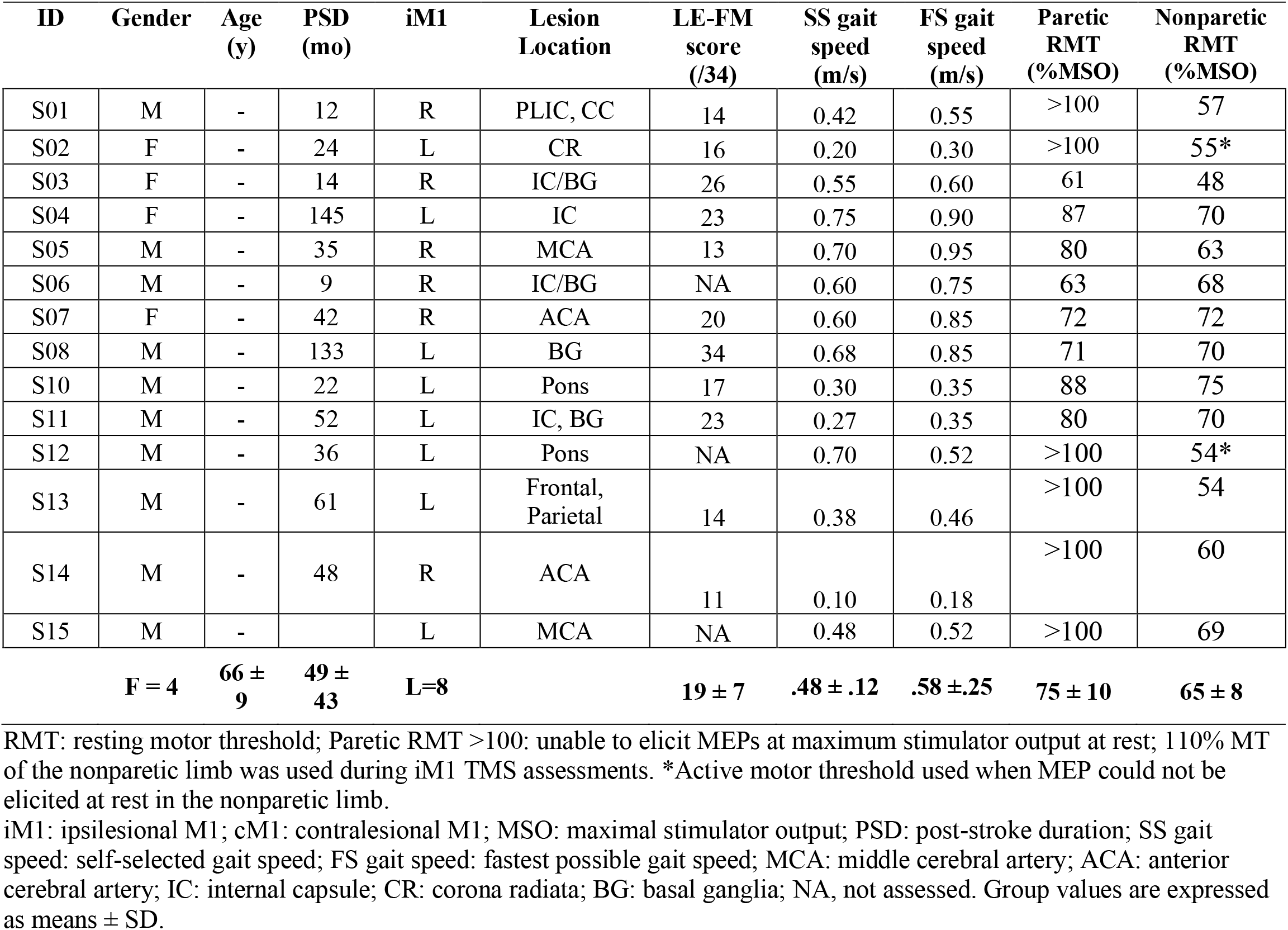
Stroke participant characteristics

### Assessment of post-stroke lower extremity clinical function and impairment

In the stroke group, clinical walking speed assessment was performed at the beginning of the neurophysiologic testing session using an overground 10-meter walk test, with participants asked to walk at their self-selected speed.^38^ An average of 3 trials was used to calculate walking speed. A subgroup of 11 stroke survivors returned for an additional testing session where additional clinical and biomechanical walking assessments were performed. During the additional session, lower extremity Fugl Myer (LE-FM) was performed to assess lower limb sensorimotor impairment.

### Assessment of biomechanical walking impairment

A biomechanical walking assessment was performed at the participant’s self-selected walking speed assessed during the overground 10-meter walk test. Kinetic and kinematic data were collected with an 8-camera motion capture system (Motion Analysis 3D Eagle, Santa Rosa, CA) while participants walked for two 30-second trials on a dual-belt treadmill (Bertec Corp., Columbus, OH, USA). Independent ground reaction forces for each limb were collected at 1000Hz from force platforms embedded within the treadmill belts.

### TMS Experimental Procedures

Prior to the first session, a high-resolution T_1_ anatomical MRI image (TR=7.4ms, TE=3.7ms, flip angle θ=6°, FOV=256mm, 160 slices, 1mm thickness) was collected for each participant. Each participant’s MRI was reconstructed using stereotactic neuronavigation software (BrainSight®, Rogue Research Inc.) and used to guide TMS delivery to the target location. Participants were fitted with earplugs and seated upright in a chair with both feet resting firmly on the ground and knees, hips and ankles positioned at ∼90 degrees. EMG activity was recorded bilaterally from TA and soleus muscles. Disposable conductive adhesive hydrogel electrodes were attached over each muscle and a ground electrode was placed over the patella of the knee following standard preparation procedures. Acqknowledge software (v. 2.2, Biopac Inc.) was used to visualize and record EMG signals using an 8-channel EMG data acquisition system.

Single monophasic TMS pulses (Magstim 200^2^, MagStim, Wales, UK) were delivered though a custom batwing coil (each wing 11cm diameter, angle between wings 65 degrees). The initial TMS coil position for targeting the lower limb M1 region was identified anatomically as 2cm lateral to the vertex of the precentral gyrus.^34,39^ The TMS coil was oriented perpendicularly to the precentral gyrus on the scalp over the target site to deliver an anterior-posterior current flow. The experimenter then adjusted the TMS coil position and orientation to most readily elicit motor-evoked potentials (MEPs) from the tibialis anterior (TA) muscle of the contralateral leg.^34^ The TA muscle was chosen to guide coil positioning because of its typically stronger corticospinal input and ease of eliciting MEPs.^33,35^ This site was used to determine the resting motor threshold (RMT) of the contralateral TA and for subsequent TMS testing procedures.^34^ If an MEP could not be elicited when the participant was at rest, then the participant was asked to lightly dorsiflex the ankle to maintain a volitional contraction at 15% of their muscle activity during a maximal force contraction.^34^ The participant maintained a low-level dorsiflexion contraction (equivalent to 15% of EMG during maximal voluntary activation) and visual feedback regarding ongoing TA EMG activation was provided while the experimenter determined the active motor threshold (AMT) for the contralateral TA. The motor threshold was determined as the TMS intensity that produced MEPs with an amplitude >50uV (resting) or >100uV (active) in at least 5 out of 10 trials.^40^ If no resting or active MEP could be produced in the paretic TA or soleus muscles (**Table 1**), the M1 site was defined as the mirror image coordinates of the contralesional (c)M1 site^41^ and 110%RMT of cM1 was used as the RMT of the ipsilesional (i)M1.^42^

Groups were matched for hemisphere of stimulation. The dominant hemisphere in controls was defined as the hemisphere contralateral to the self-reported leg used to kick a ball,^43^ and was the right leg in all control participants. TMS assessments were performed for each contralesional/dominant (c/d) M1 and ipsilesional/nondominant (i/nd)M1 during two conditions: 1) rest and 2) active ipsilateral plantarflexion contraction. During the resting condition, participants kept both feet rested on the ground. Experimenters monitored real-time EMG activity of bilateral TA and soleus muscles to ensure complete rest of bilateral lower limbs. During the active condition, participants were provided visual feedback of their soleus EMG activity while they maintained a volitional plantarflexion contraction in the leg ipsilateral to the site of TMS at 15% of soleus muscle activity during a maximal force contraction.^34^ Contraction of the limb ipsilateral to the site of TMS assessment is an established method that has been used to provide indirect assessment of transcallosal inhibition through the ipsilateral silent period.^44^ Experimenters monitored real-time EMG activity to ensure that volitional contraction levels were maintained during the experiment and that the contralateral limb remained at rest. If ipsilateral plantarflexion activity waned or if increased muscle activity was detected in the contralateral limb, the experimenter verbally cued participants to adjust. Participants were provided frequent rest breaks (∼ every 5-10 TMS pulses) to minimize fatigue or if testing conditions became compromised. A total of thirty TMS pulses was delivered at 120% RMT of the contralateral limb at a jittered rate of 0.1 to 0.25 Hz during each condition. These procedures were repeated for each limb and the order of limb and condition testing was randomized (**Figure 2**).

**Figure 2.**
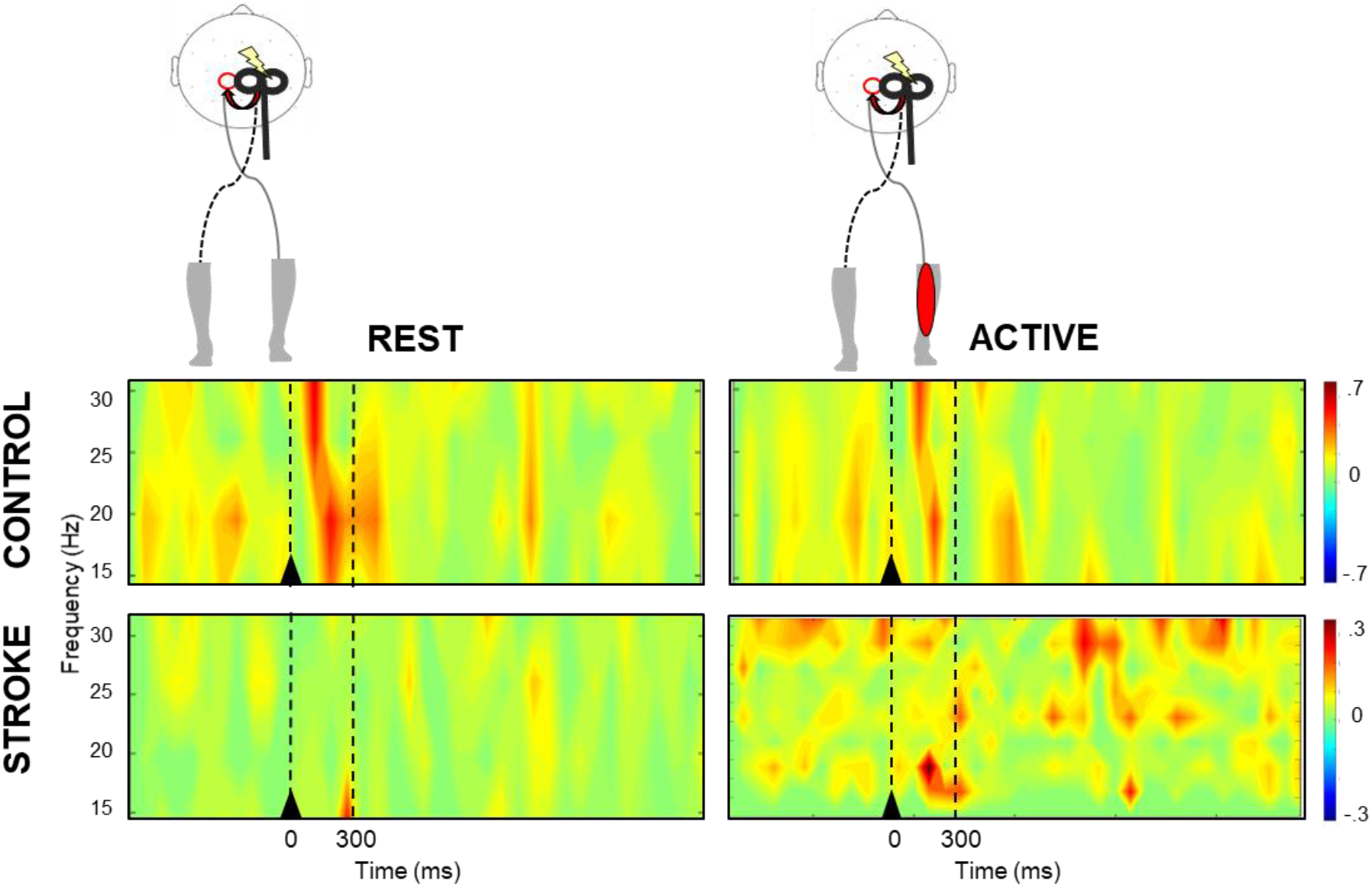
Time-frequency plots of interhemispheric cortical motor coherence during rest (left) and active (right) contralesional/nondominant M1 TMS conditions from a representative participant from the control (top) and stroke (bottom) group. During rest, the control participant showed greater TMS-evoked beta coherence compared to the active condition and stroke at rest. Black triangle denotes TMS onset. Broken lines indicate the time bin (0-300ms) of interest.

### Assessment of TMS-evoked cortical responses

Recorder software (Brain Products, GmbH) was used to record EEG signals using a 64-channel active TMS-compatible electrode cap (ActiCap, Brain Products GmbH, Gilching, Germany) connected to a BrainAmp DC amplifier (Brain Products, GmbH) during each TMS testing condition. The reference channel location was chosen as the FCz electrode position and the ground was in the AFz position. Impedance levels were lowered to ≤10 kΩ for all channels prior to TMS assessment.

### Data reduction and analysis

#### Quantification of interhemispheric coherence

EEGlab software was used to epoch (−1000 to 3000 ms relative to TMS) and re-reference (FCz electrode position) all EEG data. Imaginary part of coherence (IPC) analysis^45^ within the beta frequency range (15-30Hz) was used to calculate coherence between electrodes overlying lower limb M1 regions (left: C1, right: C2). Cortical connectivity calculated as the IPC, a methodologically conservative analyses approach, may circumvent the issue of volume conduction, or artificially inflated cortical connectivity measures due to common source signals or noise, because it requires a phase lag between sources.^45^ IPC analysis is particularly advantageous for evaluation of lower limb interhemispheric coherence due to the close anatomical proximity of lower extremity motor regions. TMS-evoked beta IPC values were calculated pre- (−300-0ms) and post- (0-300ms) TMS during rest and active conditions. Modulation of beta coherence between conditions was calculated as the difference in TMS-evoked beta coherence between active and rest conditions (active IPC – rest IPC). A value of zero indicates that beta coherence did not change; a negative value indicates that beta coherence was lower in the active condition; a positive value indicates that beta coherence was higher in the active condition.

#### Quantification of biomechanical ankle moment during gait

Kinemetic and kinetic data were filtered using a bi-directional Butterworth low-pass filter at 6 and 30 Hz. During the stance phase of each stride, the peak ankle plantarflexion moment resolved to the tibia coordinate system was calculated for the paretic and nonparetic limbs and normalized to body weight.^35^ The average peak plantarflexion ankle moment over all strides was calculated for each limb.

#### Classification of paretic leg corticospinal excitability

During the resting condition, EMG recordings of the contralateral limb during the first 100ms post-TMS were used to determine the presence or absence of a TA motor evoked potential (MEP) for each participant during i/nd M1 and c/d M1 TMS. The absence of a MEP was defined as peak-to-peak resting EMG activity <50µV in both TA and soleus muscles of the leg contralateral to the site of stimulation in > 5 out of 10 trials at 100%MSO TMS intensity.^40,46^

### Statistical analysis

Normality and homogeneity of variance of coherence, motor behavior, and biomechanics were tested using Kolmogorov-Smirnov and Levene’s tests, respectively. To test the effect of TMS on motor cortical network reactivity, we evaluated the status of cortical beta coherence at baseline (pre-TMS) and then immediately following the TMS perturbation for each group. We performed 2 separate 2×2 group(control, stroke) -by-time(pre-TMS, post-TMS) mixed-design analysis of variance (ANOVA) tests during c/d M1 and i/nd M1 TMS at rest. To test the effect of motor state on the modulation of motor cortical network connectivity, we evaluated TMS-evoked beta coherence during the rest and active conditions for each hemisphere. We performed two separate 2×2 hemisphere (i/nd M1, c/d M1) -by-condition (rest, active) within-subject ANOVA tests for each the control and stroke group. Post hoc testing was performed on significant interactions and main effects using Bonferroni corrections. We used Pearson product-moment correlation coefficients to test relationships between measures of TMS-evoked beta coherence and its modulation with motor activity versus measures of clinical impairment (LE-FM score), biomechanical impairment (paretic ankle moment), and post-stroke walking function (gait speed). We tested the association between iM1 TMS-evoked beta coherence and the presence/absence of paretic leg MEPs during the resting condition in stroke survivors in an exploratory Chi Square analysis. Statistical analyses were performed using Statistical Package for Social Sciences version 26 (IBM Corp, Armonk, NY) with an *a priori* α level set to .05.

## Results

Complete EEG datasets were collected for 8 out of 9 participants in the control group and 12 out of 14 participants in the stroke group. EEG recordings of one control participant had excessively high impedances (>50 kOhm) and were discarded from all resting and active condition analyses. One stroke survivor (S02) was unable to volitionally produce even minimal paretic plantarflexor muscle activity and was subsequently discarded from active condition analyses. One participant in the stroke group (S11) was discarded from cM1 coherence analyses due to the presence of a prolonged (>50ms) TMS artifact.

### Effect of TMS on interhemispheric beta coherence

During c/dM1 TMS during the resting condition, there was a group-by-time interaction (*F*_1,19_=4.38, *p*=.04), where beta coherence increased in response to TMS in the control (*p*=.01) but did not change in the stroke group (**Figure 2**) (*p*=.53) (**Figure 3A**). Post-TMS beta coherence in the control group was greater than stroke (*p*=.01), but no between-group differences were observed pre-TMS (*p*=.37). During i/ndM1 TMS, there was a main effect of time, where beta coherence increased following TMS, regardless of group (*F*_1,20_=6.67, *p*=.02). Though it failed to meet our adopted level of significance, there was a trend towards main effect of group, with controls showing higher beta coherence compared to stroke during iM1 TMS (*F*_1,20_=3.64, *p*=.07) (**Figure 3B**).

**Figure 3.**
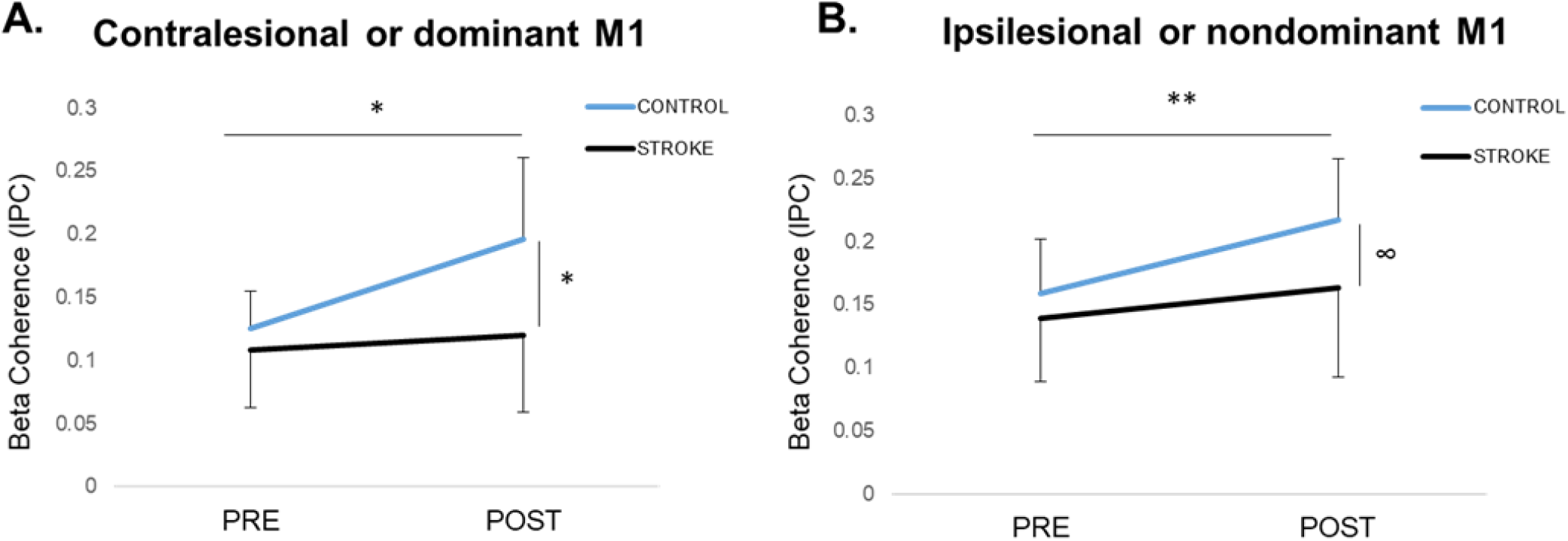
The effect of TMS on motor cortical beta coherence (mean±SD) at rest during **(A)** contralesional (c)/dominant (nd) primary motor cortex (M1) TMS and **(B)** ipsilesional (i)/nondominant (nd) M1 TMS at pre and post time points relative to the TMS. (A) During c/d M1 TMS, coherence increased from pre-to post-TMS in the control (\**p*=.01) but did not change in the stroke group (*p*=.53). There were no between-group differences pre-TMS, but controls had higher post-TMS coherence compared to stroke (\**p*=.01). (B) During i/nd M1 TMS, coherence increased from pre-to post-TMS regardless of group (\*\**p*=.02). Though group effects failed to meet our adopted level of significance, controls showed a trend of higher coherence compared to stroke (∞*p*=.07).

#### Activity-dependent modulation of TMS-evoked beta coherence

When testing the effect of active ipsilateral plantarflexor muscle contraction, the control group showed reduced TMS-evoked beta coherence during the active condition compared to rest (**Figure 4**) regardless of hemisphere of stimulation (*F*_1,8_=4.52, *p*=.03), a response that was consistent across participants (**Figure 4A**). In the stroke group, there were no differences in beta coherence between rest and active conditions (*F*_1,11_=0.08, *p*=.79), though responses were variable between participants (Figure 3B). There was a main effect of hemisphere, where beta coherence during cM1 TMS was lower compared to iM1 TMS (*F*_1,11_=4.24, *p*=.03) (**Figure 4B**).

**Figure 4.**
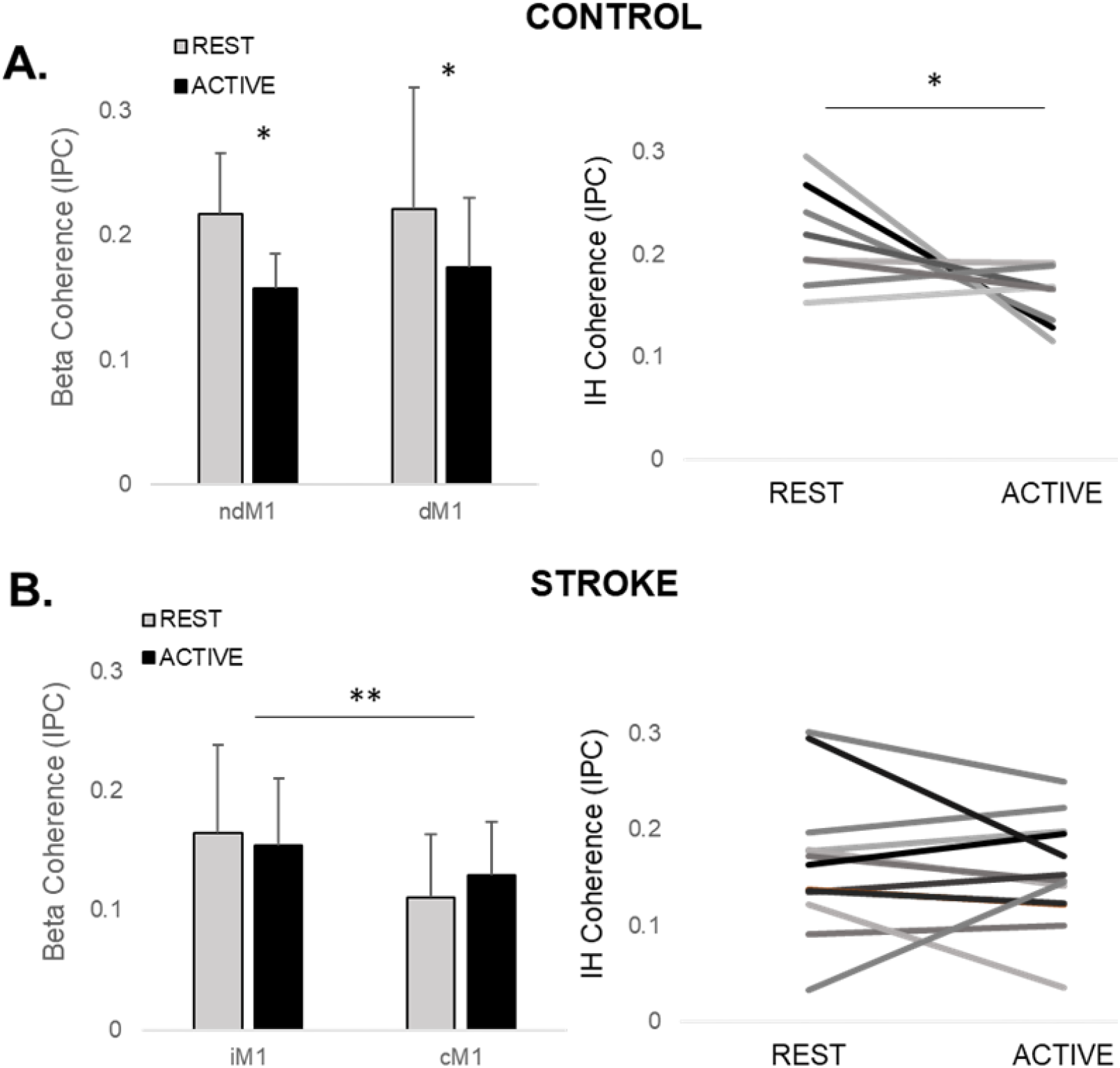
The effect of motor state on ipsilesional/nondominant (i/nd) and contralesional/dominant (c/d) M1 TMS-evoked beta coherence (mean±SD) in **(A)** controls and **(B)** stroke survivors. (A) During active ipsilateral plantarflexion, coherence consistently decreased in controls, regardless of hemisphere of stimulation (\**p*=.03). (B) In the stroke group, no differences between rest and active plantarflexion conditions were observed (p=.79 and high inter-individual variability in responses was observed In the stroke group, there was an effect of hemisphere of stimulation, where coherence during iM1 TMS was greater than cM1 TMS, regardless of condition (\*\**p*=.03).

#### Associations between TMS-evoked beta coherence and post-stroke motor behavior

Stroke survivors with higher cM1 TMS-evoked beta coherence during rest and who reduced beta coherence during paretic plantarflexion produced greater paretic limb ankle moment during gait compared to stroke survivors who had lower TMS-evoked beta coherence at rest and showed an increase from rest to active (**Figure 5A**). During rest, there was a positive relationship between cM1 TMS-evoked beta coherence and paretic ankle moment during gait (r=0.72, *p*=.03) and LE-FM (r=.68, *p*=.04) (**Figure 5B**). The modulation of TMS-evoked beta coherence from rest to active paretic plantarflexion was negatively associated with paretic ankle moment during gait (r=-0.91, *p*=.001) (Figure 4C) and LE-FM (r=-0.79, *p*=.01) (**Figure 5C**). Gait speed was not associated with TMS-evoked beta coherence at rest (r=0.52, *p*=.09) or the modulation of TMS-evoked beta coherence between rest and active conditions (r=-0.47, *p*=.12) (**Figure 5B&C**). We did not observe any relationships between iM1 TMS-evoked beta coherence at rest or during modulation from rest to nonparetic plantarflexion and motor behavior.

**Figure 5.**
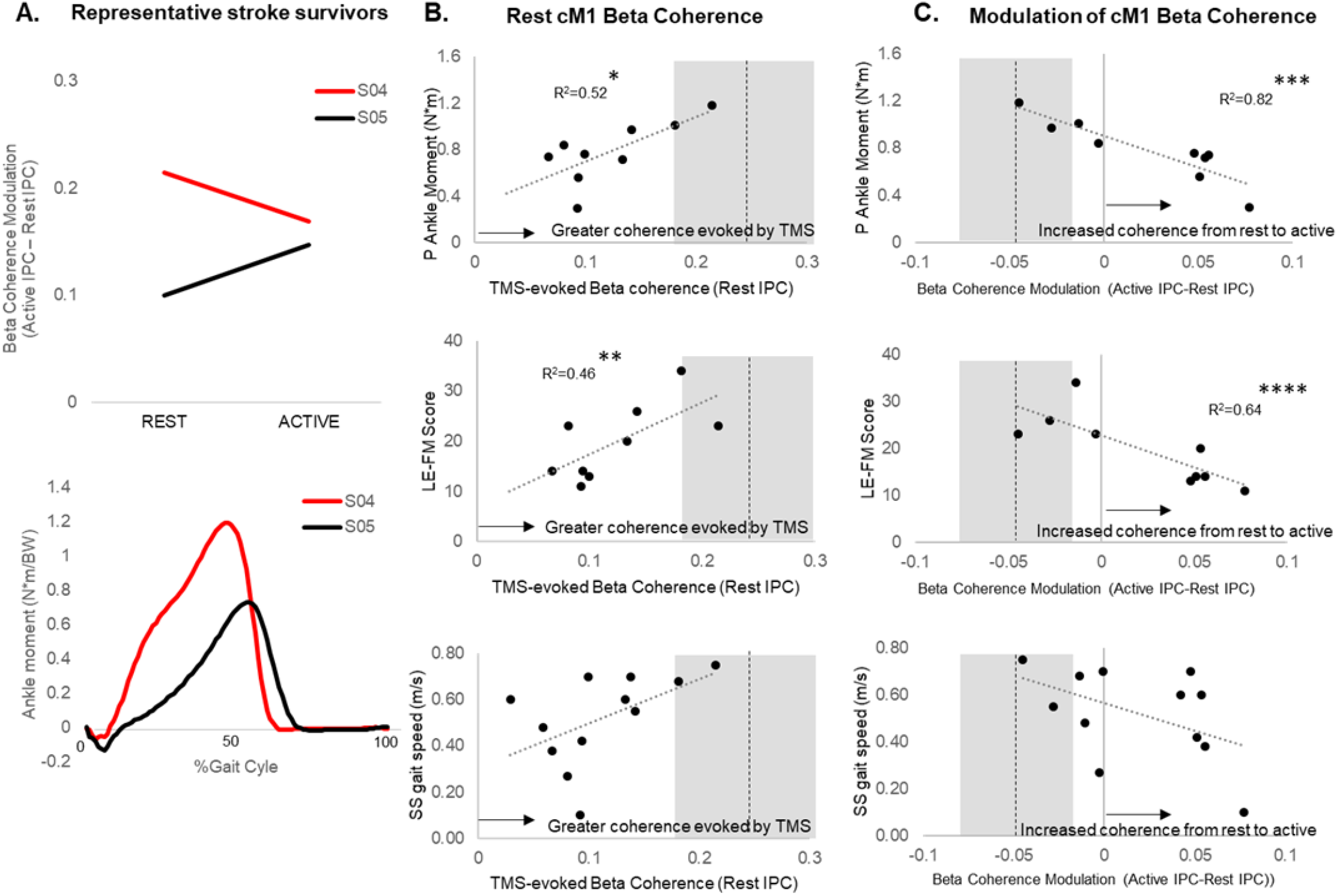
Lower limb motor behavior as a function of contralesional (c) M1 TMS-evoked beta coherence during rest and modulation of beta coherence during paretic plantarflexion. **(A)** Motor cortical beta coherence modulation from rest to active paretic plantarflexion in two representative stroke survivors with similar gait speeds (S04 and S05, see Table 1). Participant S04, showed reduced cM1 beta coherence during active paretic plantarflexion and greater paretic ankle moment during gait. Participant S05 showed increased cM1 beta coherence during active paretic plantarflexion and lower paretic ankle moment during gait. **(B)** At the group level, cM1 TMS-evoked beta coherence at rest was positively correlated with paretic ankle moment during gait (r=0.72, *p=.03) and LE-FM score (r=0.68, **p=.04) but not gait speed (r=0.52, p=.09). **(C)** Modulation of cM1 beta coherence was negatively associated with paretic ankle moment during gait (r=-0.91, ***p=.001) and LE-FM (r=-0.79, ****p=.01) but not gait speed (r=-0.47, p=.12). Broken line and grey area represent dominant M1 TMS-evoked beta coherence and modulation in controls during the same conditions (mean±SD).

#### Associations between ipsilesional motor cortical TMS-evoked beta coherence and corticospinal excitability of the paretic leg

Resting MEPs were present in the paretic limb of 8 out of 14 stroke survivors, the nonparetic limb of 12 out of 14 stroke survivors and bilaterally in 6 out of 9 controls. When MEPs were absent, this occurred in both TA and soleus muscles for all affected participants. Absent resting MEPs occurred bilaterally in 2 stroke survivors and 3 controls. Of these individuals, MEPs could be elicited with active muscle contraction in the nonparetic limb of the 2 stroke survivors and in bilateral limbs in the 2 controls. The proportion of stroke survivors with absent resting paretic TMS-evoked MEPs who showed no increase in iM1 TMS-evoked beta coherence between pre- and post-TMS time points was 0.83 (5 out of 6 participants). The proportion of stroke survivors with the presence of resting paretic MEPs who showed increased iM1 TMS-evoked beta coherence between pre- and post-TMS was 1.00 (8 out of 8 participants) (**Figure 6**). The difference in proportions of stroke survivors with absent versus present MEPs who showed increased TMS-evoked beta coherence was significant, χ^2^(1, 14) = 10.37, *p*=.001).

**Figure 6.**
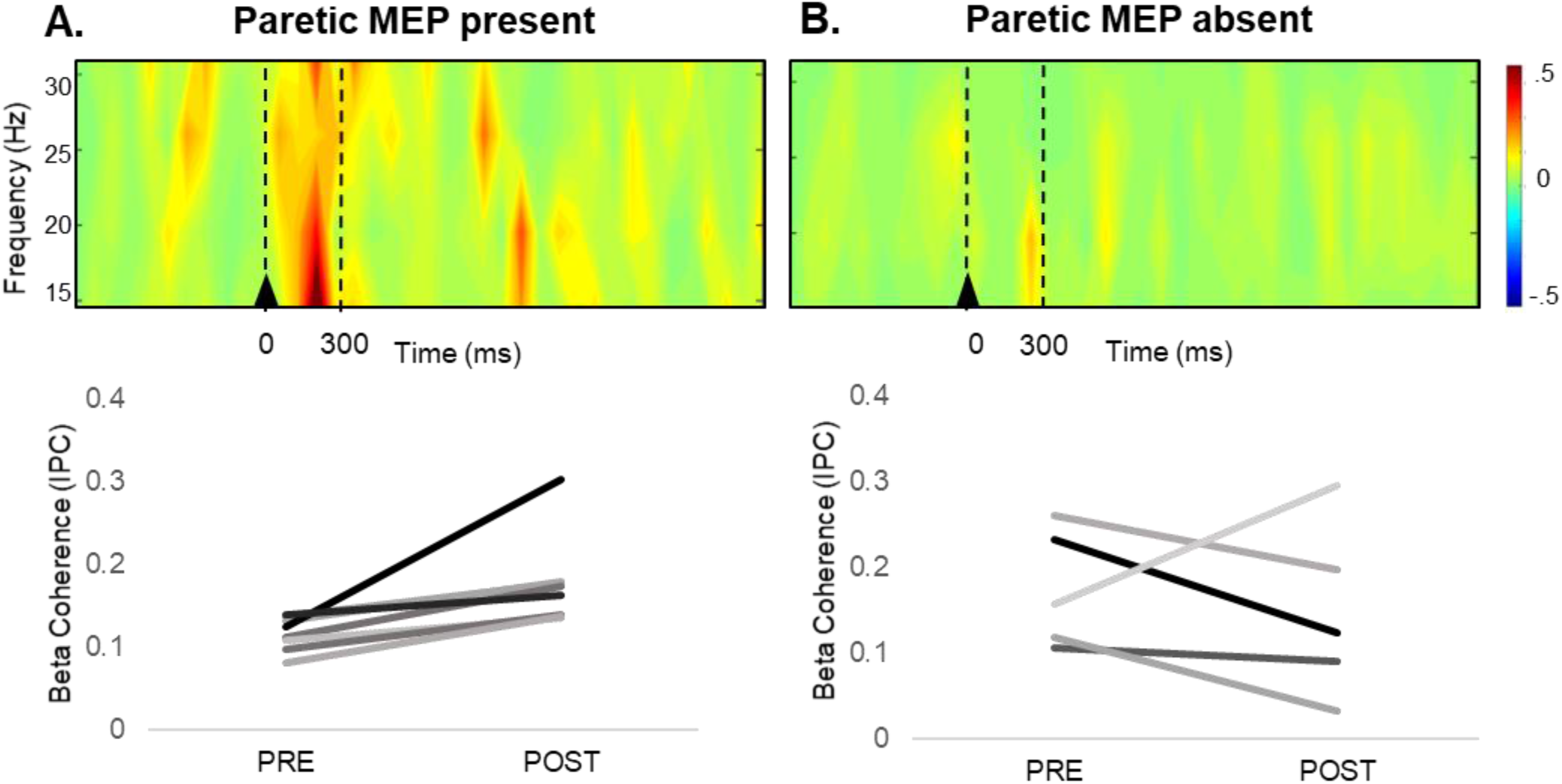
Ipsilesional (i)M1 TMS-evoked beta coherence in stroke survivors with present **(A)** and absent **(B)** motor evoked potentials (MEPs) in the paretic leg. **A)** Time-frequency plot of beta coherence in stroke survivors with present paretic leg MEPs showed increased beta coherence immediately following iM1 TMS (top). TMS-evoked increases in beta coherence during iM1 TMS were observed for all 8 stroke survivors with resting paretic leg MEPs present (bottom). **B)** Time-frequency plot of beta coherence in stroke survivors with absent paretic leg MEPs did not show a TMS-evoked increase in beta coherence (top). The lack of beta coherence increase with iM1 TMS was observed in 5 of the 6 participants with absent paretic leg MEPs (bottom). Stroke survivors with absent versus present MEPs who showed increased TMS-evoked beta coherence was significantly different χ^2^(1, 14) = 10.37, p=.001).

## Discussion

This study provides new insight into our understanding of the cortical network dynamics underpinning motor network flexibility of the lower limb corticomotor system in healthy aging and after stroke. Importantly, the present characterization of lower limb motor network flexibility builds upon previous research identifying greater cortical involvement and the presence of heightened levels of cortical inhibition during motor tasks in older adults.^47–52^ Here, we delineate the dynamic mechanisms of motor network flexibility in older adults, showing heightened motor cortical reactivity to external stimuli and reduced inhibitory network connectivity during lower limb motor activity. Our results extend these findings in neurologically-intact older adults to highlight the deficiencies of cortical motor network flexibility in stroke survivors and its link to clinical and biomechanical post-stroke walking impairment. After stroke, attenuated motor cortical reactivity to external stimuli coupled with impaired modulation of motor cortical network connectivity during lower limb motor activity were most apparent in stroke survivors with severe lower limb hemiparesis. Thus, it is unlikely that group differences in cortical motor network flexibility represent a general stroke effect, but rather reflect the reorganization of brain networks that occur over the course of post-stroke recovery. Our results also provide evidence for a novel link between ipsilesional TMS-evoked motor cortical reactivity and paretic leg corticospinal excitability (MEPs), further supporting a cortical contribution to lower limb motor network flexibility and functional mobility after stroke. These findings enhance our understanding of the normal aging lower limb corticomotor system and may have important implications for the development of individualized treatment approaches targeting specific neural deficiencies in stroke survivors, potentially maximizing post-stroke lower limb motor ability.

### Motor cortical reactivity to external stimuli and reduction after stroke

Compared to older adult controls, we found that stroke survivors showed attenuated motor cortical reactivity to external stimuli using TMS, suggesting reduced lower limb motor network flexibility within interhemispheric cortical circuits after stroke. In older adults, TMS of the primary motor cortex induced transient increases in cortical beta coherence, an effect that was absent or attenuated in stroke survivors (**Figure 3A&B**). Cortical beta coherence reflects synchrony of neural oscillations driven by inhibitory GABAergic neural neworks.^10,12,13^ After stroke, lower cortical GABA receptor density^53^ and reduced inhibitory synaptic neural activity have been observed in bilateral motor cortices.^54^ These changes could dampen the reactive capacity of cortical GABAergic networks to TMS and attenuate the typical TMS-evoked increase in cortical beta coherence. The current findings support the utility of TMS-evoked cortical responses as a potential biomarker of lower limb corticomotor system function and reactive capacity.

### Bilateral motor cortices contribute to deficits in motor network flexibility

Findings of the present study implicate the contribution of bilateral motor cortices to motor network flexibility of the lower limb corticomotor system after stroke. Attenuated motor cortical reactivity was present during stimulation of both cM1 and iM1 in stroke survivors (**Figure 3A&B**), indicating impaired motor cortical reactivity in bilateral motor cortices that occurred regardless of motor state (**Figure 4B**). Bilateral motor cortical contribution to lower limb motor network flexibility is consistent with recent neuromechanical findings by Allen et al (2019),^8^ who showed that paretic and nonparetic legs had equally impaired modulation of motor patterns across functional balance and walking tasks compared to older adult controls. Interestingly, the variability of motor pattern recruitment was *greatest in the nonparetic leg* and, in contrast to the variability of the paretic leg, was *positively associated* with post-stroke walking function.^8^ In line with these findings, in the present study we found that cortical reactivity and modulation were most impaired during cM1 stimulation (**Figure 3, Figure 4B**) and were associated with post-stroke clinical and biomechanical walking impairment (**Figure 5**). It is possible that stroke survivors who achieve the highest walking function may rely on cM1 to increase corticomotor drive to the paretic limb^33–35^ and their nonparetic leg to make flexible step-to-step adjustments in motor recruitment during walking.^8^ Together, these results may identify cortical substrates to target and modulate to potentially enhance motor network flexibility and functional mobility.

Our results provide novel evidence for differential motor cortical network reorganization mechanisms between upper and lower limbs after stroke. A greater contribution of cM1 to motor network flexibility in the lower limb motor system after stroke contrasts our previous findings in upper limb motor regions.^6,31^ Previously, we observed the greatest deficit in TMS-evoked cortical reactivity during stimulation of iM1, which was also associated with paretic upper limb motor function.^6,31^ During the process of post-stroke motor recovery, the inactivity of the paretic upper limb in contrast to the frequent, abnormal use of bilateral lower limbs during behaviors such as standing balance and walking could differentially influence motor cortical network recovery mechanisms in upper and lower limbs. Disuse of the paretic upper limb coupled with increased compensatory nonparetic limb use may contribute to a greater imbalance of upper limb iM1 and cM1 engagement, and subsequently differential motor lateralization between hemispheres.^42^ Thus, in contrast to upper limb motor regions where there is a distinct iM1 contribution to post-stroke motor network flexibility,^31^ deficits in lower limb motor network flexibly are influenced by both iM1 and cM1. Longitudinal studies tracking the trajectory of abnormal interhemispheric coherence in upper and lower limb motor regions over the course of post-stroke motor recovery will help to elucidate potential interactions between limb use, rehabilitation, and cortical motor network plasticity in ipsi- and contralesional hemispheres.

### Impaired modulation of motor network connectivity is associated with post-stroke lower limb impairment

Consistent with normal movement-related beta desynchronization that has been reported in healthy aging populations,^55^ we found reduced TMS-evoked beta coherence between motor cortices during active plantarflexion in older adult controls (**Figure 4A**). Beta desynchronization that occurs during motor activity reflects the interruption of synchronous GABAergic neural network oscillations,^10,12,13^ which are heightened in the cerebral cortex of older adults compared to their younger counterparts.^39,40^ Reduced TMS-evoked beta coherence between rest and active conditions response was remarkably consistent across older adults (**Figure 4A**). In contrast, the stroke group showed no modulation of cortical beta coherence strength between conditions, and this response was highly variable between stroke survivors (**Figure 4B**). Persistent abnormally elevated levels of synchronous beta oscillations may block or slow down changes in the “status quo” of the motor system, e.g. preventing the switch from rest to active muscle contraction.^17^ Here, our results suggest that a lack of motor network flexibility in stroke survivors manifests in the inability to disrupt synchronized GABAergic network activity between motor cortices that could contribute to greater paretic lower limb dysfunction. Supporting this notion, we found that stroke survivors with lower cortical network flexibility (lower TMS-evoked cortical reactivity and more impaired modulation of cortical beta coherence) had lower levels of clinical and biomechanical walking function (**Figure 5**). These findings are consistent with previous work showing a link between cortical oscillatory dynamics and abnormal post-stroke motor synergies during walking.^57^ Our results support the link between dynamic cortical network connectivity and post-stroke biomechanical walking impairment, further suggesting that the inability to disrupt the status quo state of motor cortical GABAergic networks (i.e. lack of motor network flexibility) may underpin pathological movement synergies that impaired paretic limb propulsive force production.

Interestingly, despite deficits in cortical motor network flexibility (e.g. reduced motor cortical reactivity to TMS (**Figure 3**), impaired modulation of motor cortical connectivity (**Figure 4**) and corticomotor excitability (e.g. the absence of paretic leg MEPs (**Table 1**)), all but one stroke survivor in this study could achieve at least some level of volitional unilateral plantarflexor muscle activation during the active condition (see Methods). In the absence of bilateral motor cortical reactivity and the ability to modulate GABAergic network connectivity that is important for normal motor activity,^17,18,55^ severely impaired stroke survivors could have potentially engaged subcortical structures to a higher degree during volitional lower limb muscle activation.^30^ In older adult populations, who demonstrate heightened reliance on the cerebral cortex for balance and walking compared to young adults,^49,51,52^ the compromised contribution of the motor cortex to lower limb motor control after stroke may be particularly detrimental to balance and walking function. Levels of intracortical inhibition in older adults has been previously associated with interlimb coordination performance during walking.^56^ Here, the reduction of TMS-evoked beta coherence that occurrs with motor activity in older adults may reflect interrupted inhibitory network connectivity from the resting motor state; activity-dependent interruption of these inhibitory motor cortical networks may serve an important functional role for the coordination of antiphasic, interlimb movement patterns necessary for walking. Here, we observed that the most severely impaired stroke survivors showed an *opposite* directional modulation of cortical beta coherence during paretic leg motor activity (**Figure 5A**). These severely impaired individuals increased cortical beta coherence during motor activity, which may reflect a neurocompensatory recruitment of transcallosal GABAergic networks that could contribute to motor interference of the contralateral leg and ineffective recruitment of paretic plantarflexor muscles. Though our assessment of cortical dynamics was performed during volitional motor activity while participants were seated, the negative relationships between cortical beta coherence modulation and paretic ankle moment during walking suggest that similar neurocompensatory mechanisms may be utilized by severely impaired stroke survivors during gait and contribute to walking dysfunction (**Figure 5C**).

### Cortical reactivity at rest yields information about functional motor network flexibility

Though information about motor network flexibility has previously been acquired primarily during the performance and execution of motor tasks,^1,16,17^ our results show that TMS-evoked cortical reactivity *at rest* can provide insight into motor network flexibility salient to post-stroke lower limb motor behavior. In mildly impaired stroke survivors, the reduction of TMS-evoked beta coherence during active paretic leg motor activity appeared to be driven by greater levels of TMS-evoked beta coherence at rest (**Figure 5A&B**). The ability to elicit greater motor cortical network reactivity at rest may therefore reflect the neural capacity for modulation of cortical network connectivity necessary during lower limb motor activity. The ability to glean information about motor network flexibility of the lower limb corticomotor system during rest could serve as a valuable prognostic tool for stroke survivors without the ability to produce volitional lower limb movement, particularly in the acute stage of stroke recovery.^58^ Notably, while we did not find statistically significant between-group differences at baseline (pre-TMS), the use of TMS to evoke cortical reactivity emboldened differences in cortical beta coherence between controls and chronic stroke survivors (**Figure 3**). These findings are in agreement with previous longitudinal assessments of resting baseline interhemispheric coherence between upper limb motor regions after stroke; in the acute stage of post-stroke recovery, interhemispheric coherence was initially elevated, but normalized over time into the chronic stage of recovery where there was no difference between stroke and controls.^58^ Future research assessing the trajectory of TMS-evoked cortical reactivity over the course of post-stroke motor recovery may provide information about the dynamics of cortical network reorganization and motor network flexibility not accessible with standalone EEG assessments.

### Absence of lower limb MEPs may be linked to impaired iM1 cortical reactivity

The degree of direct cortical influence on TMS-evoked MEPs in the lower limb has remained elusive due to the presumed strong influence of subcortical networks to lower limb motor function. For the first time, results of the present study provide novel evidence for a link between iM1 TMS-evoked cortical reactivity and paretic leg MEPs. Similar to previous studies,^24,33–35^ we were unable to elicit paretic lower limb MEPs using TMS in 43% (6 out of 14) of stroke survivors in this study. The inability to evoke a MEP has previously limited evaluation of the lower limb corticomotor system in stroke survivors who often have the most severe lower limb hemiparesis.^24^ The structural integrity of the residual corticospinal tract (CST) after stroke contributes to the ability to elicit paretic MEPs in the upper limb.^59^ However, the role of the CST in lower limb MEP production remains less clear, potentially confounded by strong subcortical network contributions to lower limb motor activity.^60^ Nonetheless, our results provide evidence that lower limb motor cortical reactivity to TMS is tightly linked to the generation of measurable TMS-evoked corticospinal output to lower limb muscles. While *all* stroke survivors with paretic lower limb MEPs (n=8) showed *increased* in cortical beta coherence immediately following iM1 TMS, 5 out of the 6 stroke survivors with absent paretic lower limb MEPs showed *no increase* in cortical beta coherence (**Figure 6**). Notably, the stroke survivor with an absent MEP but who did show an increase in iM1 cortical beta coherence (S02, **Table 1**) had a lesion that directly affected the CST. Thus, absence of a lower limb MEP may reflect both impaired TMS-evoked iM1 motor cortical reactivity and residual CST integrity after stroke, and the relationship may be driven by the lesion characteristics specific to the individual. Still, iM1 cortical reactivity and paretic corticospinal excitability did not fully explain paretic limb motor function; for example, 13 out of 14 stroke survivors were able to volitionally increase soleus muscle activity during the active condition, evidence supporting a major role of additional cortical and subcortical contributions to the production of volitional lower limb muscle contraction after stroke. Future studies utilizing multimodal neurophysiologic assessments of cortical, brainstem and spinal pathways may help to elucidate specific neural contributions to post-stroke lower limb motor recovery. This knowledge could further enable clinicians to subgroup stroke survivors based on specific neurophysiologic impairment and develop treatments targeting these individualized neurophysiologic deficits. Additionally, TMS-evoked EEG measures enabled the characterization of cortical responses from all participants, including stroke survivors with severe lower limb motor impairment and for whom no peripheral lower limb MEP responses could be elicited with TMS (**Table 1**). This capability demonstrates the utility and feasibility of TMS-evoked cortical responses in the assessment of the lower limb corticomotor system that can be used across a wide range of motor impairment levels in stroke survivors.

### Limitations

Limitations of the present study should be considered in the interpretation of the results. During TMS assessments, we carefully monitored EMG from the ipsilateral leg to ensure that no MEPs were elicited when targeting the contralateral leg; still, we cannot rule out the possibility that TMS currents could have stimulated bilateral motor cortices. As such, careful interpretation of the results involving the hemispheric directionality of motor cortical connectivity should be taken. Regardless, interhemispheric coherence would still reflect the level of functional motor cortical connectivity and comparisons across rest and active conditions would be insensitive to differential effects of bilateral M1 stimulation. Bilateral motor cortical activation is commonly observed during high effort force production.^61,62^ Here we ensured that no involuntary motor activity in the contralateral limb occurred during the active ipsilateral plantarflexion condition. Still, in severely impaired stroke survivors’ unilateral lower limb movements may present with greater effort and task complexity, and the increased interhemispheric coherence during active conditions could have been the result of bilateral motor cortical activation during higher comparable effort than unilateral motor activity. Statistical stratification of the heterogeneous stroke cohort based on lesion and demographic characteristics was not possible with the limited sample size. Though the effect of peripheral-evoked components (e.g. somatosensory and auditory potentials)^63^ may be reduced with IPC analyses,^45^ their potential contribution to TMS-evoked cortical potentials should also be acknowledged.

## Conclusions

The ability of the motor cortex to react to stimuli and modulate interhemispheric network connectivity characterizes normal lower limb motor function in older adults and appears to play an important role in the recovery of lower limb motor function after stroke. Deficits in motor network flexibility occurred bilaterally after stroke, but cortical reactivity and modulation during stimulation of the contralesional motor cortex were linked to clinical and biomechanical walking impairment of the paretic leg. Future studies that effectively target and modulate cortical origins of lower limb motor network flexibility could potentially enhance motor learning associated with post-stroke rehabilitation strategies to maximize functional mobility. By establishing TMS-EEG methodology to evaluate lower limb cortical activity, this work invites several avenues for future research utilizing TMS-evoked cortical responses to characterize the evolution of cortical network dynamics and motor network flexibility of the lower limb corticomotor system over the course of post-stroke recovery.

## Data Availability

available upon request and signing of a data sharing agreement.

## Acknowledgments and Funding

We would like to acknowledge Lewis Wheaton and Kamal Shadi for assisting in data analyses. This research was supported by the American Heart Association [AHA00035638] and the Eunice Kennedy Shriver National Institutes of Child Health & Human Development of the National Institutes of Health [F32HD096816 and K12HD055931].

